# Severe volcanic SO_2_ exposure and respiratory morbidities in the Icelandic population – a register study

**DOI:** 10.1101/19013474

**Authors:** Hanne Krage Carlsen, Unnur Valdimarsdóttir, Haraldur Briem, Francesca Dominici, Ragnhildur Gudrun Finnbjornsdottir, Thorsteinn Jóhannsson, Thor Aspelund, Thorarinn Gislason, Thorolfur Gudnason

## Abstract

**Background:** The Holuhraun volcanic eruption September 2014 to February 2015 emitted large amounts of sulfur dioxide (SO_2_).

**Objectives:** The aim of this study was to determine the association between volcanic SO_2_ gases on general population respiratory health in the Icelandic capital area some 250 km from the eruption site.

**Methods:** Respiratory health outcomes were: asthma medication dispensing (AMD) from the Icelandic Medicines Register, medical doctor consultations in primary care (PCMD) and hospital emergency department visits (HED) in Reykjavík (population: 215 000) for respiratory disease from 1 January 2010 to 31 December 2014. The associations between daily counts of health events,and daily mean SO2 concentration and high SO2 levels (24-hour mean SO2>125µg/m3) were analyzed using generalized additive models.

**Results:** After the eruption began, AMD was higher than before (129.4 *vs*. 158.4 individuals per day, *p*<0.05). Increases in SO_2_ concentration were associated with an estimated increase in AMD by 1.05% (95% CI 0.48 - 1.62%) per 10 µg/m^3^ at lag 0-2, 1.51% (95% CI 0.63 – 2.40%) increase in individuals under 18 years of age. PCMD for respiratory causes increased by 1.52% (95% CI 1.04 -2.00%) per 10 µg/m^3^ SO_2_ at lag 0-2. For HED, only effect estimated for individuals aged 64 years and older were significantly increased, by 1.541% (95% CI 0.02-3.07%) per 10 µg/m^3^ SO_2_ at lag 0-2. Following days with SO_2_ levels above 125 µg/m^3^, AMD and PCMD were increased in all age groups, in AMD mostly so in individuals under 18 by 20.4%(95%CI 4.8 – 23.4%), and adult PCMD visits by 24.1%(95%CI 16.8 – 31.3%). HED was significantly increased in elderly by 26.3% (95%CI 5.56-47.0).

**Discussion:** High levels of volcanic SO_2_ are associated with increases in dispensing of AMD, and health care utilization in primary and tertiary care. Individuals with prevalent respiratory disease may be particularly susceptible.

**Funding:** The study was funded by the Icelandic Ministry of Health.

## Introduction

The Holuhraun volcanic eruption in the Barðarbunga central volcanic system in the fall and winter of 2014-2015 was the largest eruption in Iceland since the Laki eruption in 1783-1784. Some 12 million tons of sulphur dioxide, SO_2_, was emitted from the eruption and the lava field,(Gíslason et al. 2015) and was dispersed widely over Iceland according to meteorological conditions,(Ilyinskaya et al. 2017) reaching the capital area, some 250 km from the eruption site where the 24-hour air quality guideline limit for SO_2_, 125 µg/m^3^ (WHO 2007) was exceeded repeatedly during the fall of 2014.. A previous study of professionals working at the eruption site with very high exposures experienced no serious health effects after exposure, perhaps because they were wearing protective equipment such as masks.(Carlsen et al., 2019). Meanwhile,population-level health effects remain to be investigated, in particular in sensitive populations such as children and the elderly.

SO_2_ exposure at concentrations over 500 µg/m^3^ is associated with irritation of the respiratory tract in susceptible individuals.(WHO 2007) At higher concentrations, SO_2_ exposure can trigger respiratory symptoms such as acute bronchial asthma, pulmonary oedema, and respiratory distress – especially in individuals with hyper-reactivity syndrome.(Nowak et al. 1997; WHO 2007, EPA 2008) Exposure to SO_2_ from active volcanoes is associated with increased rates chronic cough and phlegm and dry and sore throat.(Ishigami et al. 2008; Iwasawa et al. 2009, 2010; Kochi et al. 2017; Longo 2013; Longo et al. 2010, 2008; Longo and Yang 2008) Fatalities from very high SO_2_-related exposure near volcanoes have also been reported.(IVHHN.ORG) While some of the previous studies report dose-response relationship between SO_2_ and respiratory symptoms, their study designs and methods leave most of them prone to bias as symptoms were most often self-reported and participants were aware of their exposure status.(Hansell and Oppenheimer 2004) Moreover, most of the existing literature pertains to long-term area-wide exposure whereas SO_2_ exposure in Iceland during the Holuhraun eruption was intermittent with few hours or days of high SO_2_ concentrations followed by periods of low SO_2_ concentrations when wind directions changed.(Ilyinskaya et al. 2017)

With population-based registers on medicine dispensing and health care utilization as well as vigorous monitoring of air pollution in the capital area, Iceland provides an ideal setting for studying health effects of exposure to SO_2_ from volcanic eruption. The objective of this study was therefore to study the acute health effects of exposure to volcanic SO_2_ on respiratory health in the general population of Iceland. We hypothesized that respiratory health care utilization due to respiratory system illness would be increased in the general population following days with SO_2_ concentrations above the air quality guideline value of 125µg/m^3^ per 24 hours. Furthermore, we investigated the associations of health outcomes with continuous exposure, the lag-times of the effects and effects in age groups and respiratory infections and obstructive respiratory disease.

The Holuhraun volcanic eruption in North-East central Iceland began 31 August 2014 and ended 27 February 2015. The study period was 1 January 2010 – 31 December 2014, and the time before the eruption was used a reference period. The Holuhraun eruption persisted for until end of February 2015, whereas our study period ends 31 December 2014 due to a change in the database recording of events. However, SO_2_ never exceeded the air quality guideline limit during January and February 2015, although daily mean concentrations were still higher than before the eruption.

The mean population of Iceland during the study period was 320 000 inhabitants. The capital area, Reykjavík and surrounding municipalities, had 205 282 residents at the beginning of the study period, and 215 965 residents at the end.(Statistics Iceland 2017) The analysis was restricted to the capital area (residential postcodes 101-171, 200-225, and 270) where adequate information about SO_2_ exposure was available for the whole study period. The Icelandic health care system is state-centred, mainly publicly funded system with universal coverage. (Sigurgeirsdóttir et al. 2014) We obtained data on respiratory health from 1)) the National Medicines Register; 2) Primary care centres (that function as first point of contact) and 3) Landspitali, the national university hospital, the country’s centre of clinical excellence (Sigurgeirsdóttir et al. 2014). All three registers are held by the Icelandic Directorate of Health and extraction is subject to approval from the Icelandic Bioethical Committee. From the National Medicines Register we extracted data on dispensing (pharmacy sales to individuals) of prescription anti-asthma medication (AMD) classified by Anatomical Therapeutic Chemical code R03.(WHO 2015b) This class of drugs are prescribed for relieving symptoms of asthma and chronic obstructive pulmonary disease, COPD, and in some cases to relieve breathing difficulties in individuals with respiratory infections. Asthma medication dispensing is a proxy for respiratory health in a population and mild disease episodes (Menichini and Mudu, 2010; Conti et al., 2015; Furu et al., 2010). The data were extracted along with individual data on residence (postcode), age, sex and an anonymous personal identification number. From the primary care centers (PCC) and hospital emergency department (HED) databases at the Directorate of Health we extracted data on individuals diagnosed with respiratory illnesses (International Classification of Disease, ICD code J (WHO 2015a)) along with data on residence (postcode), age, sex, and an anonymous personal identification number. As the same bout of illness is likely to result in recurring health care contacts we included only the first instance of an individual registration within a 14-day period for the same diagnosis category to avoid exposure misclassification with respect to the timing of the outcome. For primary care, we analysed the number of MD visits (PCMD) in the main analysis (the total number of contacts including phone calls and consultations were analysed in a sensitivity analysis). For HED, all visits were included regardless whether the individual was admitted to hospital. For each outcome, we constructed daily time series starting 1 January 2010 to 31 December 2014 for the following age groups; children (under 18 years of age), adults (18-64 years), and elderly (age 65 years and above).

We obtained SO_2_, PM_10_, and NO_2_, data along with meteorological data from the Icelandic Environment Agency’s stationary air pollution monitor located in Reykjavík (Figure 1) for the study period and constructed a time series of daily mean values.

**Figure 1.**
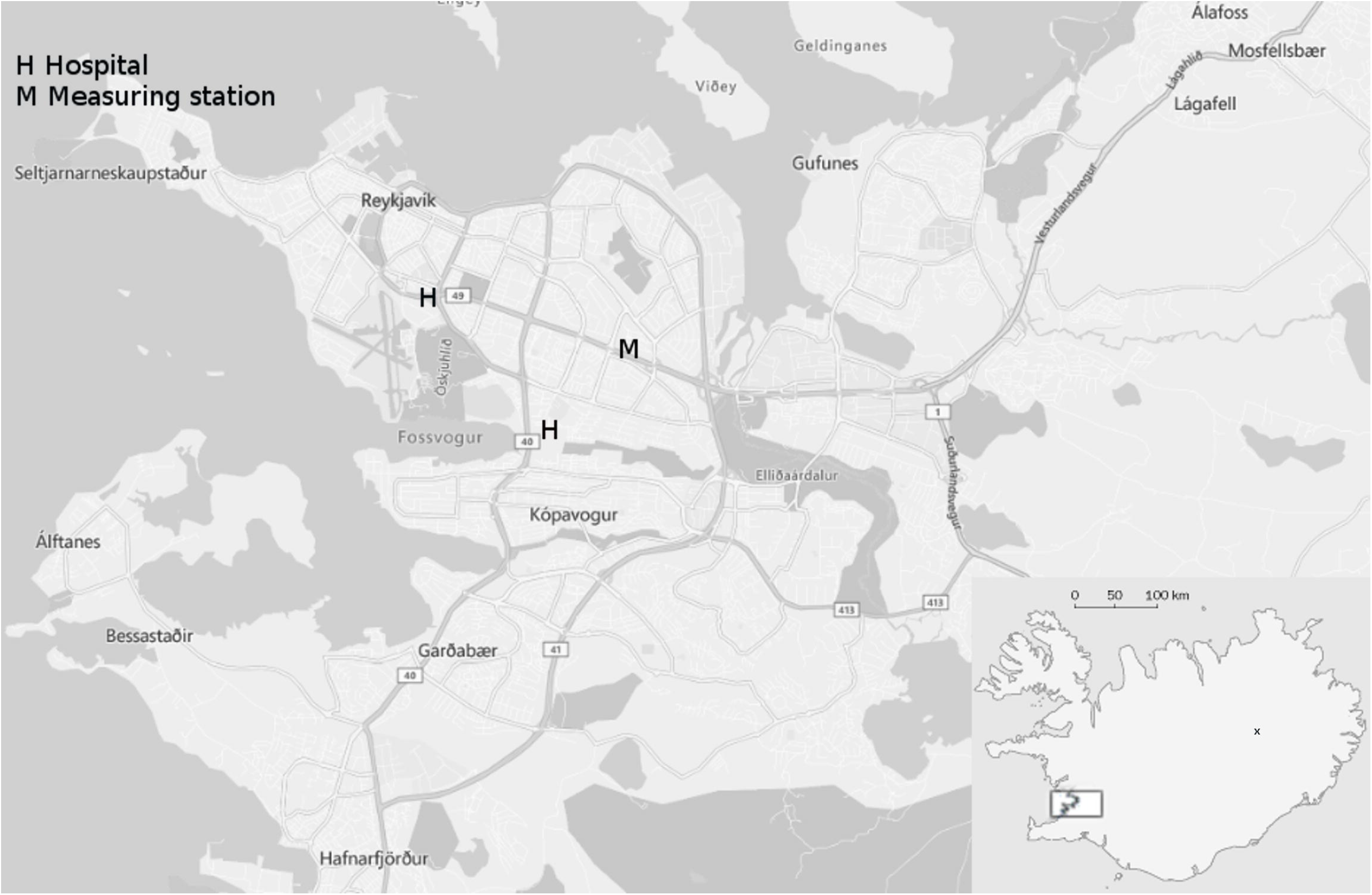
The Icelandic capital region with eruption site indicated on the inset map.

### Statistical methods

Descriptive statistics were calculated for all exposure and outcome variables for the period before and after the beginning of the eruption (using age at date of first occurrence in the data). A correlation matrix of exposure variables can be found in the supplement. We use the t-tests to assess as whether there was a statistically signficant difference in concentrations of relevant pollutant and the number of daily health outcome evens before and during the eruption.

In the regression analysis of the daily number of contacts for all outcomes during the whole study period, SO_2_ exposure was given as a) a continuous variable, or b) an indicator value of the 24-hour SO_2_ concentration exceeding the air quality guideline value (125 µg/m^3^).(WHO 2007)

We fitted to the data distributed lag non-linear models (DNLM).(Gasparrini et al. 2010) to identify the delay in days (lag days) from exposure to the observed health outcomes (Supplemental figure S1 and S2). We estimated the effects of SO_2_ exposure on the outcome by fitting generalized additive models (GAM), (Peng and Dominici, 2008)

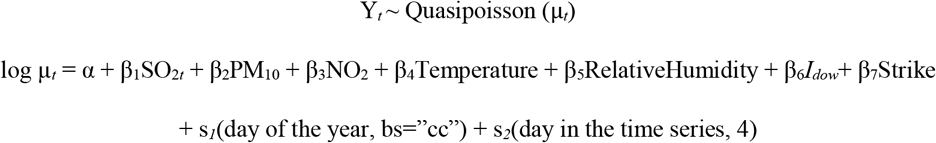

Where Y_*t*_ denotes the daily number of health events, β_1_ denotes the log relative rate of events associated with a 10 µg/m^3^ increase in SO_2_. The terms s_*1*_(day of the year, bs=“cc”) and s_*2*_(day in the time series, 4), are smoothing functions (smoothing splines) of season and calendar time designed to control for seasonal trends and trends during the whole study period, *I*_*dow*_ is an indicator for day of week and Strike is an indicator of strike days (used only in HED and PCC data)

The main exposure variable is moving three-day averages (lag 0-2) of SO_2_.We adjusted for seasonality using cubic cyclical basis, based on day of each year. We adjusted for overall time trend using smoothing spline across the entire study period with four degrees of freedom.(Bhaskaran et al. 2013) In addition, we added a factor variables to adjust for day-of week and odd holidays. For PCC and HED visits an indicator factor variable was created to adjust for days where medical doctors (MD’s) were on strike as some “strike days” were also high SO_2_ days. Several methods of addressing this issue; no adjustment, excluding strike days, or adjusting for the indicator were tested.

The results are adjusted for co-pollutants and weather (PM_10_, NO_2_, temperature, and relative humidity) at the same lag intervals as the main exposure. Results from unadjusted models are found in the supplement.

Quasipoisson distribution was assumed for all outcomes. As certain diagnoses categories may be of interest, we also present results for PCMDand HED visits stratified by subcategory into infectious respiratory disease categories for a) infectious diseases including acute upper respiratory infections (ICD codes J00-J06); influenza and pneumonia (J09-J18), and other acute lower respiratory infections (J20-J22), and b) obstructive respiratory disease, including chronic obstructive pulmonary disease, COPD, and asthma (J44 and 45).

#### Sensitivity analysis

We performed sensitivity analyses on all primary contacts including phone calls, consultations, and “other”, as actual PCMD visits are subject to availability. We also repeated the main analysis including recurring contacts for the same diagnoses within 14 days to estimate the true impact on the health care systems from increased population morbidity. For HED visits, we performed a sensitivity analysis including only individuals who were admitted for in-patient care. With regards to regular asthma medication users, official advice was broadcast to the public on days with high SO_2_ to ensure that individuals with respiratory diseases would have sufficient medicine at hand. In order to evaluate whether the observed results were merely due to compliance, we performed a sensitivity analysis excluding a) the first day, and b) the first week, in a series of days with high SO_2_ concentrations. Additionally, we performed sensitivity analysis on different lags of the HED data as our exploratory analysis revealed age-specific effects at different lags (Figure S1). To eliminate any confounding effect of other volcanic eruptions ((Eyjafjallajökull 2010 and Grímsvötn 2011) that impacted air quality during the study period,(Gudmundsson et al. 2012; Thorsteinsson et al. 2012) we repeated the main analyses for AMD, PCMD and HED excluding the years 2010 and 2011. Also, we performed analyses allowing for non-linearity of both lag structure, and concentration of SO_2_ and present those results in the supplement (Figure S1 and Figure S2) and investigated lag-responses of sub-categories of respiratory disease (Figure S3). We used an autoregressive term (adjusting for the outcome at lag 1) to improve the autocorrelation of the model residuals (Brumback et al., 2000). All analysis was performed in R using the packages “mgcv”(Wood 2018) and “dlnm”.(Gasparrini et al. 2018)

## Results

The daily mean SO_2_ concentrations in Iceland’s capital area were low or moderate, until the Holuhraun eruption began 30 August 2014 (Table 1, Figure 2, Figure S5). After the eruption began, SO_2_ concentrations surpassed the air quality guideline limit (125 µg/m^3^) on ten days, and both mean and median concentrations were significantly higher than before the eruption began.

**Table 1.**
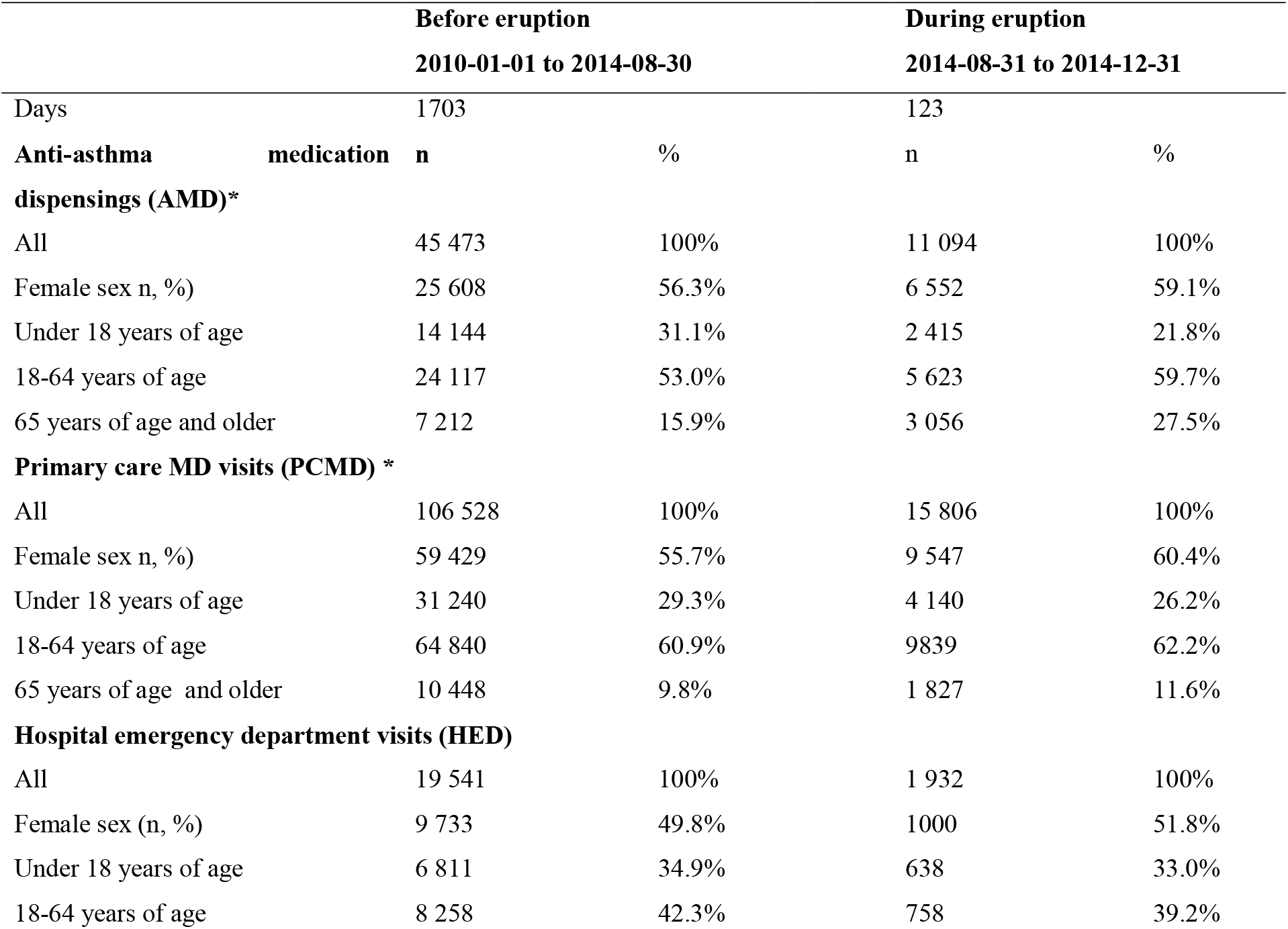
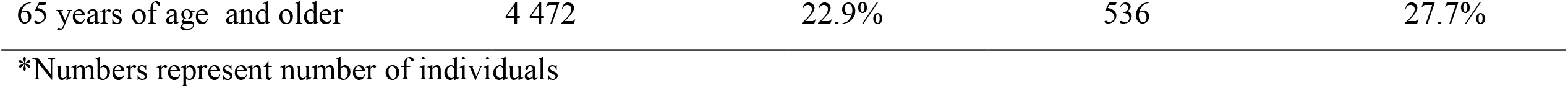
Demographic characteristics of individuals residing in the Icelandic capital who utilized health services for respiratory disease diagnoses before and during the Holuhraun eruption

**Figure 2.**
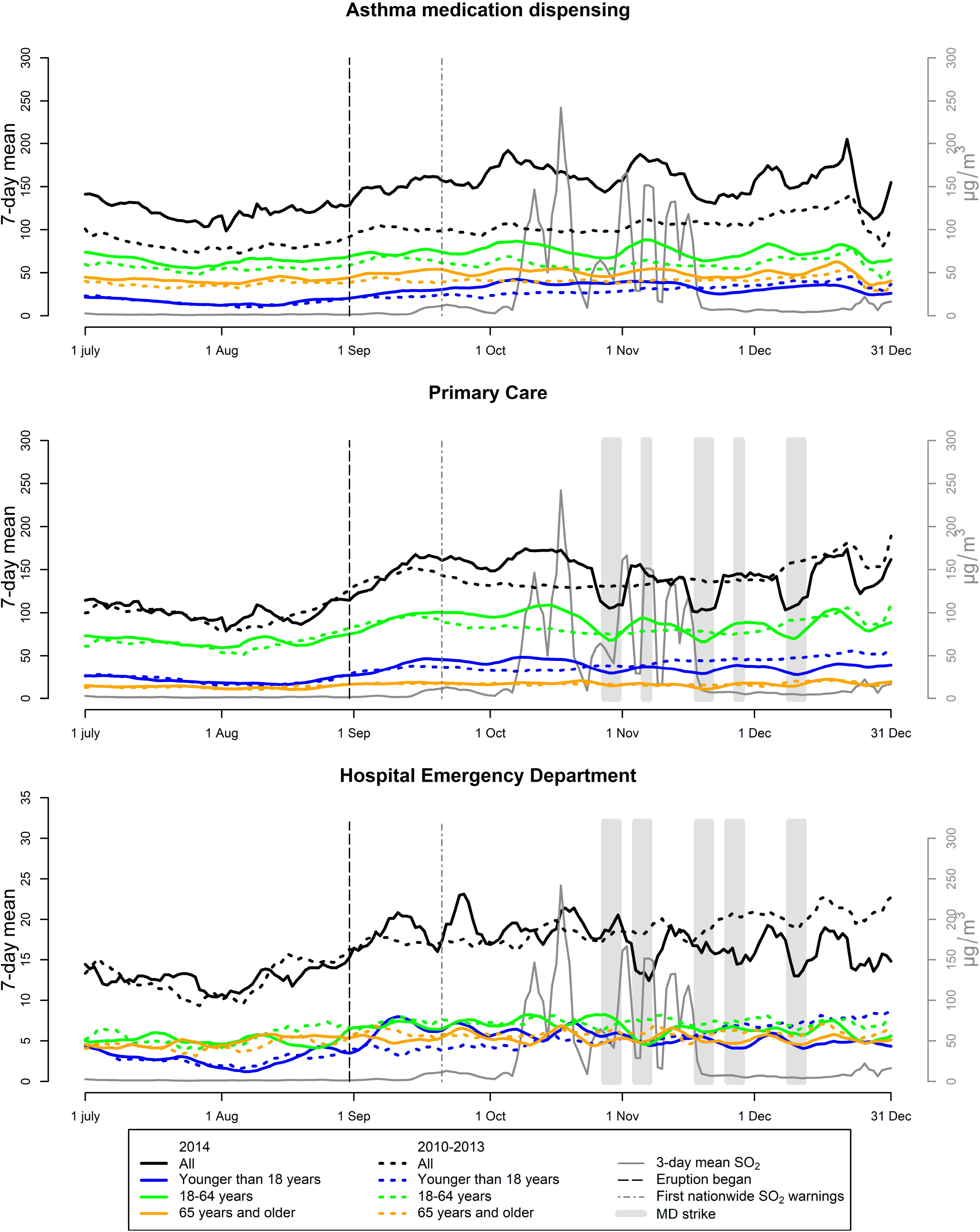
Respiratory health outcomes during the exposed period July to December 2014 (solid lines) and the unexposed previous years 2010-2013 (broken lines) for Capital area anti-asthma medication dispensing (top), primary care contacts (middle) and hospital emergency department visits (bottom) for respiratory diseases as 7-day running means. SO_2_ concentration (3-day running means) indicated in grey, broken vertical line mark the beginning of the eruption, grey vertical line denote first nation-wide SO_2_ warning. Grey shaded areas indicate MD strike in PCC and HED.

Before the eruption, an average of 129.4 individuals per day were registered purchasing (=were dispensed) AMD. The individuals, were mostly female (56.3%) and the mean age was 35.7 (SD 22.9) years. After the eruption began, there were an average of 158.4 individuals per day. The mean age was 44.5 (SD 26.3) years. Before the eruption, 106 528 individuals, attended primary care s for respiratory disease, the mean age was 32.8 (SD 22.5) years and 60.9% were women. After the eruption began, 15 806 individuals attended primary care for respiratory diseaseand the mean age was 35.0 years (SD 26.6), 60.4% were women. The most common form of contact was GP visits (61%) and phone calls (33%) (See flowchart in supplement for detail of the selection process). In HED, 19 541 individuals were registered seeking care for respiratory health outcomes before the eruption. The mean age was 36.3 years (SD 29.4) and 49.8% were women. After the eruption began, there were 1 932 individuals visited HED with a mean age of 39.4 years (SD 30.5) and 51.8% women (Table 1, Figure 2).

After the eruption began, SO_2_ was significantly higher than before; 35.7 µg/m^3^ (SD 71.2) *vs* 1.4 µg/m^3^ (SD 1.1. The daily number of number of individuals with AMD was significantly increased compared with the reference period (129.4 vs 158.4, *p*<0.001). Neither total nor age categories of HED and PCMD MD visits were significantly higher or lower during the eruption period as a whole compared to the period before (Table 2, Figure 2).

**Table 2.**
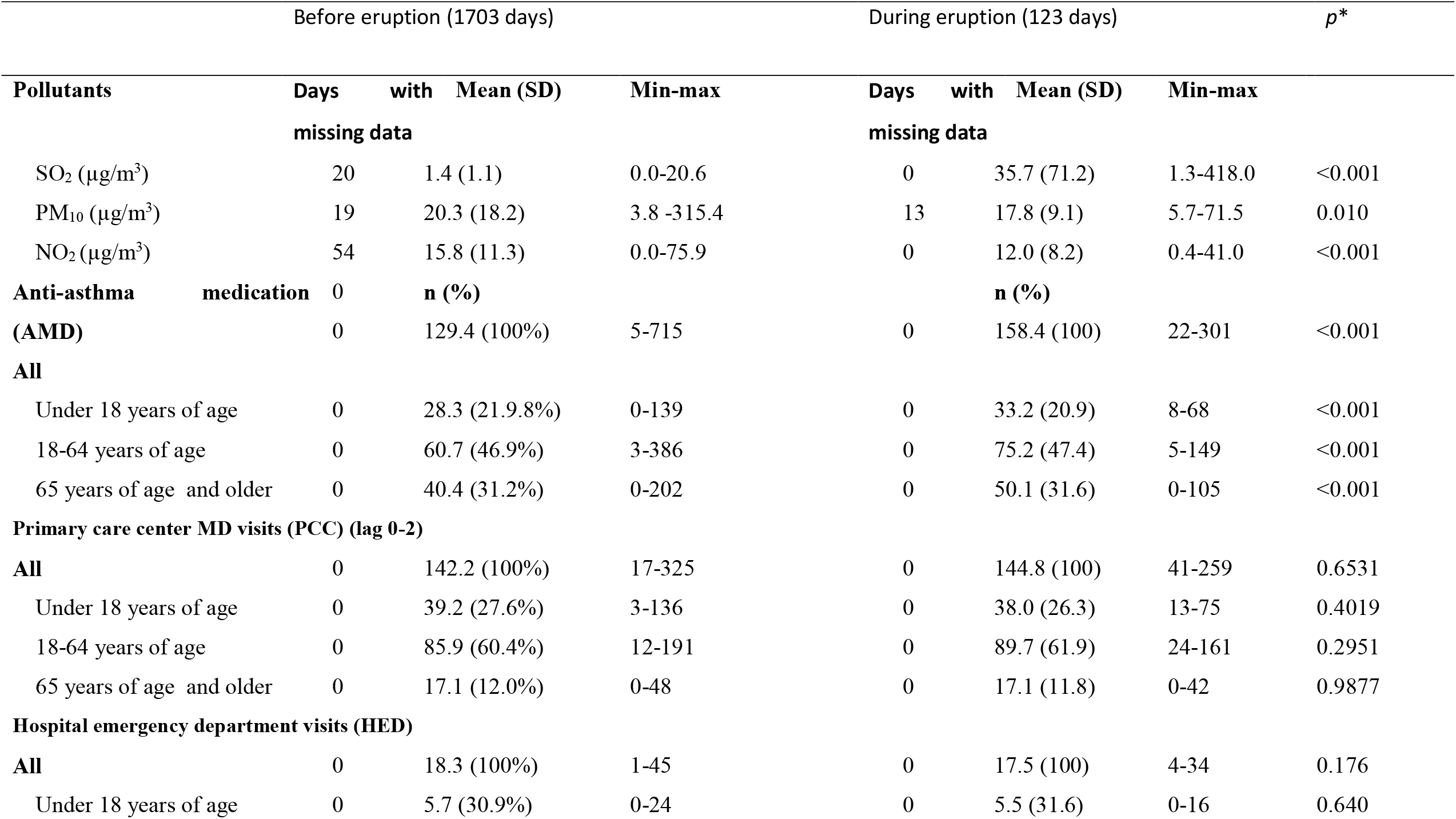
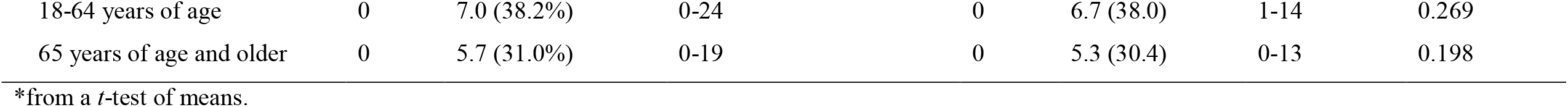
Daily pollution and respiratory health outcomes in the capital area of Iceland before and after the beginning of Holuhraun eruption.

## Regression analysis results

In the adjusted regression analyses of the whole study period 2010-2014 we found that after adjusting for other pollutants and weather, exposure to SO_2_ at short lags (lag 0-2, figure S2) was associated with an estimated increase in the number of AMDs to individuals of all ages by 1.05% (95% CI 0.48 – 1.62%) per 10 µg/m^3^ SO_2_. For children 0-17 years of age, the estimate was higher, 1.51% (95% CI 0.63 – 2.40%) and for individuals 65 years and older, the estimate was lower, 0.77% (95% CI 0.05 – 1.51). SO_2_ concentrations exceeding the air quality guideline of 125µg/m^3^, were associated with a statistically significant increase in dispensing of AMD at lag 0–2 by 13.5% (95% CI 5.3-21.7%), the increase was 20.4% (95% CI 8.0-32.9%) in individuals under 18 years of age (Table 3). In the age-stratified analysis, the association effect estimates were highest in children under 18 by 20.4% (95% CI 8.0 - 32.9%) and with a lower estimate for elderly, 8.8% (95%CI -1.9 – 19.5%) which did not reach statistical significance.

**Table 3.** Percent excess risk associated with daily SO_2_ exposure at lag 0-2 (both as a continuous variable and as an indicator for days with pollution levels above the air quality guideline value) and changes in respiratory health outcomes in the capital area of Iceland

|  | SO<sub>2</sub> (per 10 µg/m<sup>3</sup>) |  | SO<sub>2</sub> levels >125 µg/m<sup>3</sup> |  |
| --- | --- | --- | --- | --- |
|  | % | 95% CI | % | 95% CI |
| **Asthma medication (AMD)** |  |  |  |  |
| All | 1.05% | 0.48 – 1.62% | 13.5% | 5.3 – 21.7% |
| Children 0-17 | 1.51% | 0.63 – 2.40% | 20.4% | 8.0 – 32.9% |
| Adults 18-65 | 1.01% | 0.37 – 1.66% | 14.1% | 4.8 – 23.4% |
| Elderly >65 | 0.77% | 0.05 – 1.51% | 8.8% | -1.9 – 19.5% |
| **Primary care MD visits (PCMD)** |  |  |  |  |
| All | 1.52% | 1.04 – 2.00% | 21.6% | 15.1 – 28.6% |
| Children 0-17 | 1.31% | 0.53 – 2.10% | 15.3% | 4.4 – 26.2% |
| Adults 18-65 | 1.52% | 1.01 – 2.04% | 24.1% | 16.8 – 31.3% |
| Elderly >65 | 1.36% | 0.44 – 2.28% | 15.7% | 2.4 – 29.9% |
| **Hospital Emergency Department (HED)** |  |  |  |  |
| All | 1.12% | 0.19 – 2.05% | 11.2% | -1.64 – 24.0% |
| Children 0-17 | 1.02% | -0.68 – 2.74% | 5.4% | -19.14 – 30.0% |
| Adults 18-65 | 1.18% | -0.25 – 2.63% | 6.8% | -13.44 – 27.1% |
| Elderly >65 | 1.54% | 0.02 – 3.07% | 26.3% | 5.56 – 47.0% |

All models are adjusted for season, time trend, day of week, odd holidays, the outcome at lag 1, temperature, relative humidity, NO<sub>2</sub>, PM<sub>10</sub> at the same lags as SO<sub>2</sub>. n=1705.

For PCMD and HED, including an indicator variable for days where MDs were on strike improved the model fit and was an effect modifier which increased the effect estimates, so this variable was added to the PCMD and HED models. PCMD visits increased by 1.52% (95% CI 1.04 – 2.00%) per 10 µg/m^3^ SO_2_ at lag 0–2 for all ages, and stratifying by age, the increase was highest in adults.

For PCMD, short term exposure to SO_2_ over the air quality guideline value was associated with increases by 21.6% (95% CI 15.1 – 28.6%). The increase was highest in adults by 24.1% (95% CI 16.8 – 31.3%).

For HED visits, SO_2_ exposure was associated with increased risk for individuals older than 64 at lag 0– 2 (Figure S1) by 1.54% (95% CI 0.02 – 3.07%) per 10 µg/m^3^ (Table 3). For exposure over the air quality guideline limit, the effect estimate significantly increased only in elderly by 26.3% (95% CI 5.56 – 47.0%).

Stratifying by diagnosis category, SO_2_ exposure was associated with increased PCMD visits for respiratory infections at lag 0–2 by 1.35% (95% CI 0.80 – 1.89%) per 10 µg/m^3^. PCC contacts for obstructive disease were increased by 2.48% (95% CI 1.57 – 3.39%). Following air quality guideline value exceedances, the estimated increase in PCC MD visits were 19.0 (95% CI 10.5 – 28.1%) and 34.3% (95% CI 18.5 – 52.2%) for respiratory infections and obstructive disease, respectively. For HED visits, SO2 exposure was not associated with significant increases in visits due to respiratory infections or asthma or COPD although all estimates were positive (Table 4).

**Table 4.** Percent excess risk associated with daily SO_2_ exposure (both as a continuous variable and as an indicator for days with pollution levels above the air quality guideline value) and changes in respiratory health in diagnosis subcategories in primary care and hospital emergency departments in the capital area of Iceland

|  | Mean (SD) | SO<sub>2</sub> (per 10 µg/m<sup>3</sup>)* |  | SO<sub>2</sub> levels >125 SO<sub>2</sub> ug/m<sup>3</sup> |  |
| --- | --- | --- | --- | --- | --- |
|  |  | % | 95% CI | % | 95% CI |
| Primary care MD visits (PCMD) (lag 0-2) † |  |  |  |  |  |
| Respiratory infections † | 108.6 (46.2) | 1.35% | 0.80 – 1.89% | 19.0% | 10.5 – 28.1% |
| Asthma and COPD (J44-45) | 15.6 (9.6) | 2.48% | 1.57 – 3.39% | 34.3% | 18.5 – 52.2% |
| Hospital emergency department visits (HED) (lag 0-2) |  |  |  |  |  |
| Respiratory infections † | 9.3 (4.3) | 1.29% | 0.07 – 2.51% | 11.2% | -43.7 – 31.0% |
| Asthma and COPD (J44-45) | 4.3 (2.7) | 0.90% | -0.82 – 2.64% | 4.0% | -17.9 – 31.8% |

\* Adjusted for season (spline), time trend (spline), day of week, odd holidays, the outcome at lag 1, temperature, relative humidity, NO<sub>2</sub>, PM<sub>10</sub> at the same lags as SO<sub>2</sub>. † ICD codes starting with J0x, J1x, and J2x.

In the sensitivity analysis, the unadjusted associations between SO_2_ exposure and health outcomes was only marginally different (<10 percentage point change) from the adjusted results (Table S1) with the exception of the estimates for HED visits which decreased from 1.12% (95% CI 0.19 – 11.2%) to 0.84% (95% CI -0.08-1.76%). However, the estimated association with HED visits in elderly increased from 1.54% (95% CI 0.02 – 3.70%) in the adjusted analysis to 1.70% (0.26 -3.16%) in the unadjusted analysis.

### Sensitivity analysis

In sensitivity analyses of the outcome variables, considering all PCC contacts including phone calls and consultations, the estimated effect of SO_2_ increased slightly, most so in elderly, where the estimate increased from 1.36 (95% CI 0.44 - 2.28%) per 10 µg/m^3^ SO_2_ in the main analysis (Table 3) to 1.44% (95% CI 0.69 – 2.19%) (Table S2). Not excluding the recurring contacts (individuals with two or more PCC contacts within 14 days), the age distribution had shifted further, so the estimated increase in events in elderly reached 1.56% (95% CI 0.85 – 2.28%) per 10 µg/m^3^. The estimated effects of exposure to SO_2_ concentrations above the air quality guidelines on AMD and PCMD followed a similar pattern to that of continuous exposure where adults had the highest effect estimated. In the analysis of HED admissions rather than all visits, all effect estimates were positive, but only HED visits in children at lag 2-4 reached statistical significance and were estimated to increase 6.0% (95% CI 1.29-10.7%) per 10 µg/m^3^ SO_2_ (Table S2) and 175.2% (95% CI 20.9-526.2%) after SO_2_ levels over the air quality guideline limit. In this analysis, there were no significant associations between SO_2_ and HED at lag 0-2.

In sensitivity analysis to investigate the importance of compliance on the observed associations with AMD and PCC, we removed the first day in a series of days with high SO_2_ concentrations (Table S3). The estimated effect of SO_2_ exposure on AMD and PCC outcomes was higher for children than in the main analysis, but lower in adults compared to the main analysis. Excluding the whole week after the first high SO_2_ concentrations in the capital area, effect estimates for both AMD and PCC were higher in elderly and adults compared to the main analysis. Considering effects on HED of SO_2_ exposure at lag 2-4 altered the age-distribution of the results, so that there HED in children were increased by 2.09% (95% CI 0.51-3.70%)

Excluding the years 2010 and 2011 where ash-rich volcanic eruptions occurred yielded generally similar, albeit slightly attenuated estimates which in few cases failed to reach statistical significance. Also, the association between HED and SO_2_ at lag 2-4 were calculated (Table S3).

## Discussion

In this study, a comparison of health care utilisation in Iceland’s capital are before and after the eruption began showed that only AMD increased significantly during the Holuhraun eruption, but primary care and hospital care was not higher during the eruption than before. However, in adjusted time series regression analysis, SO_2_ concentrations were associated with increased AMD, MD visits in primary care for respiratory causes and hospital emergency department visitsat lag 0-2. Using a cut-off or continuous measure of SO_2_ resulted in largely similar estimates (Table 3). The effect estimates for specific disease categories indicate that individuals with obstructive lung disease were particularly affected (Table 4).

In the regression, our choice of lags were informed - exploratory analysis (Figures S1). For AMD and PCC visits the observed increase occurred during the same day and up to two days after a peak in SO_2_ concentration. HED visits increased one to three days after a peak in SO_2_ which could indicate that primary care is the first point of contact for a majority of individuals and hospital care is sought only after other care options have been exhausted. In the compliance analysis, the effect of initial warnings of volcanic gas exposure appeared to be limited, as excluding the first day or week of high SO_2_ yielded similar estimates for AMD and PCC visits, although there was some loss of statistical power. This indicates that the increase reflected actual increased respiratory morbidities rather than only compliance as it persisted after first exposure (Supplemental Table S2). We present both unadjusted results (Table S1) and results adjusted for other air pollutants (Tables 3 and 4) as well as plotted an exposure-response function with a spline. Comparing the results from the continuous analysis and the cut-off analysis, we found little indication of significant non-linear effects, which is also observed in the spline plot of the exposure-response function (Figure S2). Our results are mostly consistent with those from previous studies of SO_2_ exposure in urban settings; e.g. respiratory mortality rates were estimated to increase by 2.4% per 27 µg/m^3^ SO_2_ (Li et al. 2017), corresponding to a point estimate of 0.88% per 10 µg/m^3^ SO_2_, which is lower than the estimates for HED and PCMD estimate of respiratory morbidity associated with SO_2_ found in this study; 1.12% (95% CI 0.19-2.05%) and 1.52% (95% CI 1.04 – 2.00%) per 10 µg/m^3^ SO_2_, respectively. In previous studies of SO_2_ exposure during volcanic eruptions, the SO_2_ concentration on Miyakejima Island increased after Mount Oyama erupted in 2000. After evacuating the island at the time of the eruption, residents and aid workers returned from 2005 and onwards. Aid volunteers (n=611) reported respiratory irritation and the rates were associated with SO_2_ exposure - particularly among women and non-smokers.(Ishigami et al. 2008) During the same eruption, children, who were exposed to daily mean concentration of 125 µg/m^3^ SO_2_, had increased rates of wheezing.(Iwasawa et al. 2010) In a follow-up study 2006-2011, permanent residents of Miyakejima Island (n=168) who lived in areas with SO_2_ reported increased rates of cough and wheeze few effects.(Kochi et al. 2017) None of the studies of people exposed at Miyakejima Island found adverse effects on lung function.(Ishigami et al. 2008; Iwasawa et al. 2009, 2010; Kochi et al. 2017) The Kilauea Volcano in Hawaii has been erupting continuously since 1983 with periods of increased eruption activity from 2008. SO_2_ concentrations spiked from a daily mean concentration of 72 µg/m^3^ to 215 µg/m^3^ in a populated area downwind during this period and the rates of cough, and acute pharyngitis diagnosed at a clinic in the exposed community increased subsequently.(Longo et al. 2010) A cross sectional study found that those living in areas exposed to SO_2_ had increased risk of cough, phlegm, and hay fever. A qualitative survey of subjects from the same study population revealed that participants reported that symptoms disappeared 7-10 days after leaving the exposed area, but reappeared one week after returning.(Longo et al. 2008) In a study of individuals exposed at the Holuhraun eruption site we found that while some reported respiratory symptoms during their stay at the eruption site, most symptoms had resolved and lung function was normal at a clinical examination one to six days later.(Carlsen et al. 2019) Regarding individual susceptibility, we observed the highest effect estimates for PCMD visits for asthma and COPD (Table 4). The effect estimate for HED visits were also increased but did not reach statistical significance. In controlled trials, hyper-responsiveness to SO_2_ has previously been reported as common (20-25%) in individuals with positive asthma test (methacoline test).(Nowak et al. 1997), indicating that they be particularly vulnerable to severe SO_2_ exposure. Similarly, respiratory infections were increased in PCMD visits and tended to be increased for HED visits, although the increase did not reach statistical significance. Finally, an association between SO_2_ and hospital visits for upper respiratory tract infections has previously been reported(Li et al. 2017).

Previous studies of health effects of air pollution in Iceland have yielded lower effect estimates of daily air pollution on morbidity than the current study,(Carlsen et al. 2012b) other air pollution exposure types are thus not likely to bias the results. This includes H_2_S, which has a low correlation (<0.1) with the exposure of interest during the study period (data not shown). During the reference period, there were two ash-rich volcanic eruptions, in Eyjafjallajökull 2010 and Grímsvötn 2011. While ash from Eyjafjallajökull had local respiratory health effects(Carlsen et al. 2012a) and there may have been adverse health effects in the capital area,(Carlsen et al. 2015) neither eruption had significant SO_2_ emissions and sensitivity analysis excluding this period yielded similar results (Supplemental Table S2). Our study employs a time series design were individual level risk factors are not time-dependent and thus should not confound the association between short term exposure to air SO_2_ and health outcomes, leaving bias due to unmeasured confounders, seasonal variation, and other intermittent exposures as main concerns. The exposed period October and November of 2014 did not coincide with any viral respiratory illnesses (e.g. influenza and RS-virus) epidemic,(Tilkynningarskyldir sjúkdómar 2018) but it did coincide with the MD labour conflict which resulted in lower PCC and HED attendance during those days, Hence, results from the analysis comparing the eruption period with the time before may have underestimated of true effect of the SO_2_ from the eruption.

SO_2_, PM_10_ and NO_2_ data was missing for a number of days which were excluded from the analysis. As the volcanic plume effectively changed the chemical composition of the atmosphere during the eruption period (Supplemental Tables S4a-b), the correlations of SO_2_ with other air pollutants were altered after the eruption (we present unadjusted results in the supplement (Supplemental Table S1)). The results from these are nearly identical to the main analysis (Table 3), indicating that the effect of the very high concentrations of SO_2_ was unambiguous.

It is a strength of the current study that health data were collected prospectively from population-wide registers, which minimizes the risk of information bias from individuals knowing their exposure status. Although the exposure would have been known to the public for at least part of the exposed period but we attempt to addresse this source of bias in the sensitivity analysis, showing only moderate changes to the results (Table S3). As a study outcome, we use dispensing of asthma-medication, a novel and more sensitive proxy than primary care attendance and hospital visits. It measures morbidity in individuals who in most cases already are in contact with the health care system. However, this outcome has been used in other studies and is known to be a proxy for asthma morbidity in a population (Furu et al,. 2010; Naureckas et al., 2005) and be associated with air pollution (Menichini and Mudu, 2010; Fattore et al., 2016) A limitation with register-data is that it is not collected for research purposes and diagnoses given in the health care system could be biased towards overestimation as medical professionals assume respiratory outcomes to be more likely during the eruption. However, as the estimated effect are similar across all respiratory outcomes we conclude that this sources of bias is not likely to fully explain our findings.. By basing exposure status on residential postcode we assume that people spend most of their time at home, however, outcomes in individuals not in the exposed area during the study period would bias the results toward the null. Reykjavík and the Icelandic capital area was exposed to SO_2_ concentrations above 125µg/m^3^ during a total of ten days which occurred mostly during October and November 2014. This limits the statistical power of the study and our options for further analysis, as does the fact that our study period did not extend until after the eruption, meaning that we cannot fully assess the importance of population increase during the study period on our results.

In conclusion, this comprehensive study with prospectively collected data on volcanic air pollution exposure and respiratory outcome, is the first to firmly establish an association between spikes of high SO_2_ concentrations and respiratory outcomes in the general population, particularly in individuals with prevalent respiratory disease. These findings emphasise the need for attention from authorities and parents to the health susceptible individuals during times of volcanic eruptions.

## Data Availability

The full health data contains sensitive individual-level information and is not publicly available. The health data can be made available to researchers after approval of a formal application to the Icelandic Directorate of Health and the Icelandic Bioethics committee (reference: VSNb2015050022/03.01). The data from the EAI air quality monitoring stations is publicly available in real-time, and archived data can be obtained from the EAI upon request.

## Acknowledgements

The authors wish to acknowledge the The Environment Agency of Iceland and Kristinn Jónsson from the Icelandic Directorate of health for providing data, and Dr. Evgenya Ilyinskaya for valuable comments on the manuscript. The study was funded by the Icelandic Ministry of Health. The funding source had no influence on the reporting or publication of the study.

